# Self-reported chronotypes are associated with greater time-of-day differences in substrate oxidation following morning and evening mixed-meal challenges

**DOI:** 10.64898/2026.11.19.26368014

**Authors:** Eleanor U. Flacke, Nicholas A. Foreman, Stevaughn Smith, Eden Glick, Angela Nguyen, Adam Ciarleglio, Lauren N. Woodie, Matthew D. Barberio

**Affiliations:** Department of Exercise and Nutrition Science, The Milken Institute School of Public Health, George Washington University, Washington, D.C; Department of Biostatistics and Bioinformatics, The Milken Institute School of Public Health, George Washington University, Washington, D.C; Department of Pharmacology & Physiology, School of Medicine and Health Sciences, George Washington University, Washington, D.C

**Keywords:** Circadian rhythm, substrate oxidation, metabolism, fat oxidation, chrononutrition

## Abstract

**BACKGROUND:** Postprandial metabolic and vascular responses are associated with insulin sensitivity, which displays diurnal rhythmicity. How meal timing interacts with chronotype to affect substrate utilization and vascular response to feeding is unclear.

**METHODS:** In a randomized crossover trial, 18 otherwise healthy young adults (9 females; ages 18–45 y/o) completed two isocaloric mixed-meal challenges (MMCs) at 08:30 or 17:00 preceded by a 10-hour fast. Before and for two hours after each MMC, expired gases via canopy indirect calorimetry were used to calculate respiratory exchange ratio (RER) and metabolic flexibility (ΔRER). We measured central and brachial blood pressures via pulse wave analysis and arterial stiffness via carotid-femoral pulse wave velocity (cf-PWV) at 0, 60,120 minutes. The Composite Morningness Questionnaire (CMQ) measured chronotype. We used functional regression and linear mixed-effects models to analyze metabolic and hemodynamic outcomes, respectively.

**RESULTS:** During fasting (p=0.043) and 0-90 minutes postprandial (p=0.001), evening RER was lower than morning RER without differences in metabolic flexibility. In the evening, morning chronotypes have lower fasting RER, while evening chronotypes show a delayed increase in postprandial RER compared to other chronotypes. Intermediate chronotypes have a similar postprandial RER response independent of time of day. There was no effect of meal condition on augmentation index normalized to 75 bpm (p=0.310) or arterial stiffness (p=0.882).

**CONCLUSION:** Despite lower evening RER, postprandial metabolic flexibility is similar between morning and evening meals. More extreme chronotypes are associated with greater differences in substrate oxidation between morning and evening meals without differences in fasting or postprandial arterial stiffness.

**New & Noteworthy:** Circadian rhythms influence insulin sensitivity and therefore may change metabolic flexibility to the same morning and evening meal. We show greater fat oxidation to an evening meal but no difference in metabolic flexibility in healthy young adults. We extend these findings by showing more extreme self-reported chronotypes have greater morning-to-evening differences in substrate oxidation despite similar metabolic flexibility.

## Introduction

Circadian rhythms are orchestrated by the suprachiasmatic nucleus (SCN), which synchronizes multiple organ systems with daylight to prime physiology for food intake during the day (active phase) and fasting during the night (inactive phase) [1, 2]. The SCN and peripheral clocks of the pancreas, liver, and adipose tissues drive downstream diurnal variation in insulin sensitivity [3], lipolysis, and hepatic metabolism [4], thereby altering nutrient handling across the 24-hour day [5]. Simultaneously, diurnal variation in catecholamines and autonomic activity are thought to drive observed circadian rhythmicity in blood pressure [6] and heart rate [7, 8]. In response to mixed macronutrient feeding, previous work has observed decreases in metabolic flexibility (i.e., the shift in substrate oxidation from fasting to feeding), and increases in postprandial fat oxidation, across the course of the day (i.e., breakfast to lunch to dinner) [9]. Given the decreases in sympathetic vascular tone [10] [8] and increases in postprandial glycemia and insulinemia [11] from morning to evening, postprandial vasodilation and consequent hemodynamic responses to feeding may also change from morning to evening.

Therefore, the same metabolic stimulus, a meal, can elicit wide-ranging cardiometabolic responses depending on time of day. Shift work, which alters the timing of behavioral stimuli (i.e., sleep, feeding, light exposure), is associated with greater cardiovascular disease risk [12]. Similarly, short-term extreme acute experimental desynchronization decouples central and peripheral clocks [13] and increases postprandial plasma glucose, insulin, and mean arterial pressure [14]. Even a shift in the distribution of caloric intake from earlier to later in the day is associated with higher BMI and worse insulin sensitivity [15]. One potential mechanism for these deleterious effects is blunted metabolic flexibility (i.e., the shift from fasting to feeding) due to a reduction in insulin sensitivity throughout the day [3], as greater insulin sensitivity is associated with greater metabolic flexibility [16], even in otherwise healthy young adults [17]. It is unknown how metabolic flexibility differs from morning to evening following an isocaloric meal.

“Chronotype”, which refers to the phenotypic expression of endogenous rhythms, varies depending on genetics, long-term behavioral patterns (e.g. shift work), geography, age, and sex [18, 19]. Meta-analysis of observational studies between chronotype and higher risk of type 2 diabetes, higher HbA1C, and CVD-related mortality highlight considerable between-study heterogeneity [20–22], underscoring the need for further research accounting for chronotype.

Importantly, few experimental studies have considered temporal preference for habitual activity, and most questionnaire-based outpatient assessment methods for chronotype are treated as dichotomous or categorical variables, which is incongruent with the actual distribution of circadian rhythm and chronotype [19]. The influence of chronotype on changes to postprandial metabolism across the day remains unclear. Additionally, work that has considered the effect of chronotype on responses to morning vs evening feeding has not simultaneously assessed cardiovascular outcomes [23]. To properly understand time of day differences in physiological responses to feeding, we must study these inter-related systems in concert.

In a sample of otherwise healthy young adults without overt pathology, we tested the effect of time-of-day on substrate oxidation, postprandial metabolic flexibility, arterial stiffness, and central and peripheral hemodynamics in response to an isocaloric mixed-macronutrient liquid meal challenge. We defined metabolic flexibility as the change from the fasted to fed states, using functional regression [24] to leverage our continuous indirect calorimetry. We further explored the association between self-reported chronotype, as measured by preference for morning activity via the Composite Morningness Questionnaire [25], and postprandial substrate oxidation in the morning and evening. We hypothesized that metabolic flexibility to a mixed-meal challenge (MMC) would be reduced (i.e., lower ΔRER) in the evening (17:00) versus the morning (08:30) following a 10-hour fast, and that later self-reported activity would modify the substrate oxidation response to an MMC in the morning *vs* evening. We also hypothesized arterial stiffness would decrease from fasting to the postprandial state with greater decreases following evening feeding than morning feeding.

## Methods

Young adults (ages 18-45 y/o; BMI: 18.5-40.0 kg/m^2^; n=18) without overt pathology were recruited from October 2024 to January 2026 via paper and electronic flyers posted around the George Washington University (GWU) campus (Washington, DC, USA). Participants were eligible if they were deemed low risk for medical complications during exercise [26] and denied taking medications known to impact metabolic outcomes, including antioxidant supplements, antibiotics, steroids, probiotics, exogenous sex hormones other than hormonal anti-contraceptives, or medications for management of cholesterol, diabetes, or obesity. Participants with a history of disordered eating, gastrointestinal diseases, food allergies, habitual consumption of high-fat or ketogenic diets, or recent food insecurity [27] were also excluded. Screening criteria and documentation of informed consent [28] were conducted electronically via the Research Electronic Data Capture (REDCap) platform prior to the first laboratory visit [29, 30]. We screened 249 participants, of which 112 were eligible to participate and 36 chose to enroll in the study. All study procedures were approved by the Institutional Review Board at The George Washington University (NCR246197) and registered on ClinicalTrials.gov (NCT07122765). Of the 36 enrolled participants, we present results from the 18 participants (9 males, 9 females) who completed both morning and evening MMCs.

### Experimental Design

Participants completed three laboratory visits each with ≥ 48 hours between visits (**Figure 1**). For each participant, we assessed body composition and maximal aerobic capacity before completing two MMCs in a randomized, crossover design. The MMCs followed measurement of fasting, resting energy expenditure (REE). Visits started at 08:30 and 17:00, respectively, with MMC consumption 45-60 minutes in. Assessments of body composition and aerobic capacity were performed between 08:00 and 15:00. For participants randomized to complete their first MMC in the evening, we also measured resting metabolic rate prior to the graded exercise test. Participants also completed questionnaires documenting race, ethnicity, medical history, and medication use. Standard questionnaires were used to assess habitual physical activity [31], habitual sleep quality [32], and habitual diet as defined by the alignment between habitual diet and the Mediterranean Diet [33] and the percentage of calories from fat [34]. Notably, we used the CMQ to assess self-reported preference for morning activity, where a higher score indicates a greater preference for morning activities [25]. All research visits were conducted in the GWU Metabolism and Exercise Testing laboratory.

**Figure 1:**
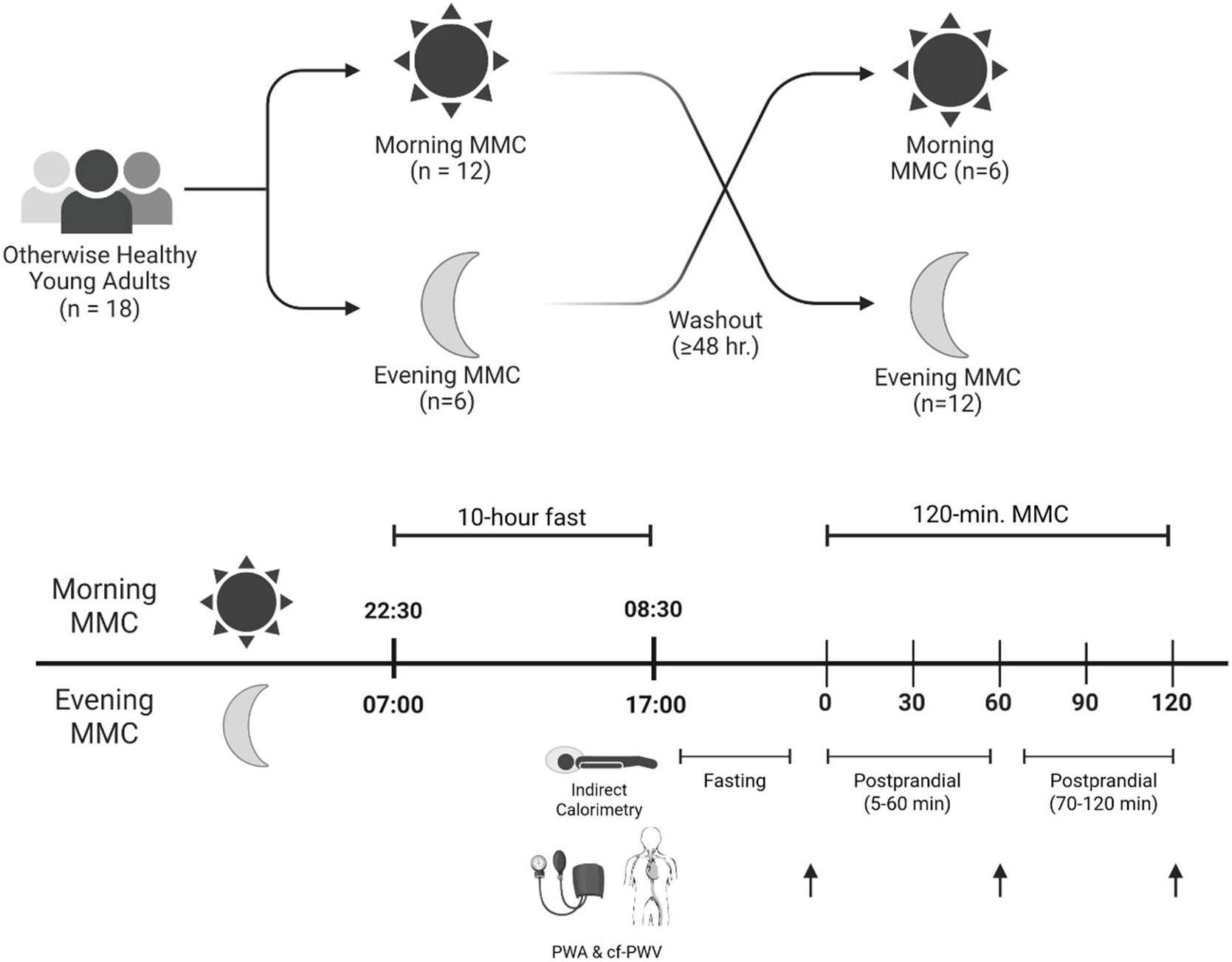
Experimental Design: Figure 1 caption: Participants (n=18) completed two MMCs each in this crossover trial: one in the morning and evening, each preceded by a 10-hour fast. MMC dose was adjusted to give 25% of estimated TDEE. Prior to MMC consumption, participants completed fasting indirect calorimetry, carotid-femoral pulse wave velocity, and central and brachial blood pressure measurements; these measures were repeated at the indicated timepoints through 120 minutes postprandial. Figure created in BioRender.

### Baseline Assessments

Prior to assessment of body composition and aerobic capacity, participants were instructed to fast for at least four hours, to avoid strenuous exercise or consuming alcohol, nicotine, or tobacco for 24 hours, and to avoid caffeine for 12 hours. Upon arrival, hydration was measured using a portable refractometer. If urine specific gravity was above 1.020, participants were given 230 mL (∼8 oz.) of water. Height was measured using a wall-mounted stadiometer (Seca 216; Hamburg, Germany) to the nearest 0.1 cm. Weight was measured using the digital scale as part of the bioelectrical impedance analysis (InBody 770; Seoul, South Korea). Dual X-ray absorptiometry (GE Lunar iDXA; GE Healthcare, Chicago, IL) with manufacturer software (enCORE Forma version 18) was used to measure body fat percentage (BF%), fat-free mass (FFM), fat mass, and visceral adipose tissue (VAT) volume. Waist circumference was measured as the narrowest circumference between the iliac crest and the bottom rib and repeated until duplicate measures were within 0.5 cm [35]. For all body composition analyses, participants removed all jewelry and wore clothing free of metal zippers or reflective surfaces. A cycling graded exercise test was conducted to assess cardiorespiratory fitness (see *Supplementary Materials*).

### Metabolic Control

Prior to each MMC, participants were instructed to refrain from structured exercise (e.g., any movement for health benefits such as running, sports, resistance training, etc.) or consuming alcohol, nicotine, or tobacco for 24 hours and caffeine for 12 hours. Each MMC was conducted after a ten-hour fast. For the evening MMC, participants were instructed to consume a large breakfast 10-11 hours prior to the scheduled start of the evening MMC. Adherence to these stipulations was confirmed at the start of each MMC.

### Mixed Meal Challenge

At the start of each MMC, body weight was measured via electronic scale (see *Baseline Assessments*). For 20-25 minutes, participants lay supine on a padded table in a dimly lit room, and indirect calorimetry was conducted via the canopy method (Parvo Medics TrueOne 2400; Salt Lake City, UT); this data is represented as time 0 in subsequent analyses and figures. On the right arm, we measured blood pressure in duplicate before using pulse wave analysis (PWA) via Sphygmocor XCEL (CONNEQT Health; Naperville, IL) to measure the central blood pressure waveform. PWA was repeated if the manufacturer’s quality control metric was not ≥ 80%. Manufacturer software also applied wave separation methods to calculate the amplitude of forward (Pf) and backward (Pb) pressure [36]. Arterial stiffness was measured via carotid-femoral pulse wave velocity (cf-PWV) [37].

Participants completed indirect calorimetry to estimate REE after an overnight fast. This was performed either at the screening visit prior to exercise or immediately prior to the first MMC, depending on equipment availability. We created rolling five-minute averages of REE, excluding the first ten minutes of data, and selected the lowest REE which also had a coefficient of variation for VO_2_ and VCO_2_ ≤10% [38]. Because there is only consensus for how to process indirect calorimetry data in rodents [39] and not in humans, we utilized this method to ensure stable data for calculation of REE while also acknowledging that lower energy expenditure is likely closer to a true resting value. We estimated sedentary TDEE as 1.3 * REE [40].

Participants consumed the meal shake (Ensure Plus; Abbott; Abbott Park, Illinois; 55% carbohydrates, 28% fat, 18% protein) in five minutes or less, followed by up to 150 mL of water if desired. MMC dose was individualized to provide 25% of REE-estimated TDEE. We denoted the start of the postprandial period as the first sip of the meal shake. From five to 60 and from 70 to 120 minutes postprandial, we conducted indirect calorimetry via the canopy method. To mitigate moisture build-up from prolonged indirect calorimetry, we replaced the viral filter and tubing into the mixing chamber at 60 minutes postprandial. Blood pressure, PWA, and cf-PWV measurements were repeated at 60 and 120 minutes postprandial.

### Data Preparation

We calculated energy expenditure (EE; kcal/min) and rates of fat (FOX; kcal/min) and carbohydrate (CHOX; kcal/min) oxidation using published equations [41, 42] assuming negligible urinary nitrogen excretion. Following the application of a six-minute rolling average and outlier removal on postprandial substrate metabolism data, we interpolated each outcome measure into one-minute intervals [43], allowing us to satisfy the requirement for complete data with functional regression [24]. Further data processing details are provided in the *Supplemental Materials*.

## Statistical Analysis

We used mixed effects models to determine the effect of meal condition on our cardiovascular outcomes. For each outcome, we treated time as a three-level factor with a condition-by-time interaction, the corresponding main effects, and a random intercept for participant to account for repeated measures. In the absence of a significant interaction, we tested a main effects model adjusted for sex with a random intercept for participant. If necessary, outcome measures were transformed via natural log after visual review of model assumptions via the performance package (version 0.15.1) [44]. We used the emmeans package [45] for pairwise comparisons with a Tukey adjustment. For significant main effects, we report mean difference ± SE with Satterthwaite 95% confidence intervals (CI).

We used functional regression to determine the effect of meal condition on substrate oxidation. This approach models an intensively sampled outcome variable as a function over time rather than individual outcome measures at discrete timepoints [24]. Compared to a conventional ANOVA or mixed models approach, functional regression limits the Type I error rate associated with significance testing the across 120 sampled timepoints. Specifically, we used a function-on-scalar regression model estimated via a penalized fitting approach [46] employed by the pffr function from the R refund package (version 0.1-40) [47]. We fit functional regression models for each whole-body metabolic outcome after centering to mean zero and standard deviation of one, using the mean and SD in our sample. Each model includes binary predictors for meal condition, male sex, and a subject-specific random intercept function to account for repeated functional outcome measures. For analyses of metabolic flexibility, we further adjusted for the fasting value obtained immediately prior to the MMC (i.e., always after a 10-hour fast). To visualize our results, we present raw data for each outcome as mean ± SD for each condition along with panels showing each pairwise difference across conditions. Colored shading shows where the 95% CI does not cross zero, indicating a significant effect of meal condition on the outcome at the shaded timepoint(s).

We further analyzed the effect of chronotype on postprandial RER. Because it was an exploratory outcome, we simplified our analyses of chronotype to RER in order to simplify interpretation across levels of CMQ. We defined chronotype using the CMQ score as a continuous variable to capture the spread of behavior throughout the day [19]. We fit a model with a chronotype*meal condition interaction and compared it to a corresponding main effects model using an F-test. We further explored a potential nonlinear effect of chronotype by comparing it against the same model with a linear effect of chronotype. We display modeled data as mean ± SE of morning and evening EE, RER, FOX, and CHOX at three representative CMQ values in our sample: 42 as mean + 1 SD; 34 as mean; and 26 as mean - 1 SD. All analyses were conducted in R (version 4.4.2). We present raw data as mean ± SD and statistical estimates in-text as mean ± SE. We interpret statistical significance using an *a priori* alpha level of 0.05 or a 95% CI excluding zero.

## Results

### Participants

Participants (n=18, 9 female) were young adults (21.8 ± 3.4 years) without any previously diagnosed cardiometabolic pathology (Table 1). On average, participants were overweight by BMI (25.2 ± 3.5 kg/m^2^) but spanned the BMI spectrum of normal weight (6 males, 5 females), overweight (2 males, 3 females), or obesity (1 male, 1 female). Using a CMQ score of ≥ 44 or ≤ 22 to denote morning and evening chronotypes from intermediate [25], participants were predominantly intermediate (n=16) with 2 morning (range: 23-49). Participants consumed 430 ± 80 mL of the mixed-meal shake, providing 635 ± 120 kcal (see *Mixed Meal Challenge*).

**Table 1.** Participant Characteristics.

|  | Overall (n=18) | Female (n=9) | Male (n=9) |
|---|---|---|---|
| Age (years) | 22 (3) | 21 (2) | 22 (4) |
| Height (cm) | 172.0 (8.5) | 166.8 (6.9) | 177.3 (6.8) |
| Weight (kg) | 74.6 (12.5) | 70.0 (8.3) | 79.2 (14.7) |
| BMI (kg/m<sup>2</sup>) | 25.2 (3.5) | 25.2 (3.6) | 25.1 (3.7) |
| Body fat (%) | 30.7 (8.5) | 35.1 (6.5) | 26.3 (8.3) |
| Subcutaneous fat mass (lbs) | 3.1 (1.7) | 3.2 (1.4) | 3.1 (2.1) |
| VAT mass (lbs) | 1.0 (0.9) | 0.6 (0.7) | 1.4 (0.9) |
| Resting HR (bpm) | 61 (11) | 63 (10) | 59 (11) |
| Brachial SBP (mmHg) | 115 (8) | 113 (6) | 116 (9) |
| Brachial DBP (mmHg) | 66 (8) | 66 (8) | 66 (9) |
| Central SBP (mmHg) | 101 (7) | 100 (6) | 103 (7) |
| Central DBP (mmHg) | 67 (8) | 67 (8) | 66 (9) |
| Brachial MAP (mmHg) | 80 (8) | 80 (8) | 80 (9) |
| cf-PWV (m/s) | 5.3 (0.9) | 4.9 (0.6) | 5.8 (0.9) |
| VO<sub>2PEAK</sub> (mL/kg/min) | 32.9 (7.0) | 31.3 (8.1) | 35.0 (5.0) |
| VO<sub>2PEAK</sub> (percentile) [48] | 39 (21) | 46 (24) | 28 (8) |
| CMQ | 34 (8) | 35 (10) | 34 (6) |

Data are mean (SD). Body mass index (BMI); Carotid-femoral pulse wave velocity (cf-PWV); Composite Morningness-eveningness Questionnaire (CMQ); Diastolic blood pressure (DBP); Heart rate (HR); Mean arterial pressure (MAP); Systolic blood pressure (SBP); Visceral adipose tissue (VAT)

### Energy Expenditure and Substrate Oxidation

Fasting EE was not different between morning and evening (mean ± SD: 1.12 ± 0.16 vs. 1.16 ± 0.2 kcal/min; p=0.171 via paired t-test). In the evening, fasting RER was lower (0.82 ± 0.05 vs. 0.78 ± 0.06; p=0.043) and FOX was higher (0.64 ± 0.0.19 vs. 0.83 ± 0.26 kcal/min; p=0.011) without differences in CHOX (0.48 ± 0.24 vs. 0.32 ± 0.29 kcal/min; p=0.084) (**Figure 2**). In functional regression models, there was a significant effect of meal condition on postprandial RER (p=0.001; mean difference ± SD: −0.04 ± 0.005), FOX (p<0.001; 0.07 ± 0.01 kcal/min), and CHOX (p<0.001; −0.17 ± 0.02 kcal/min). Review of 95% confidence intervals from functional regression models shows evening RER was lower through 90 minutes postprandial with higher FOX and lower CHOX compared to the morning. After 90 minutes postprandial, there were no differences in RER, FOX, or CHOX (**Figure 2A-C**). While there was an overall effect of meal condition on postprandial EE (p = 0.004), there were no significant differences between morning and evening EE at any one timepoint. Equipment availability limited our ability to obtain the resting EE measurement for all participants prior to the randomization of visit order; this measurement was necessary to prescribe the MMC dose for the evening visit in participants where the evening visit was the first visit. As a result, more participants completed the morning MMC before the evening visit (n=12) than vice versa (n=6).

**Figure 2.**
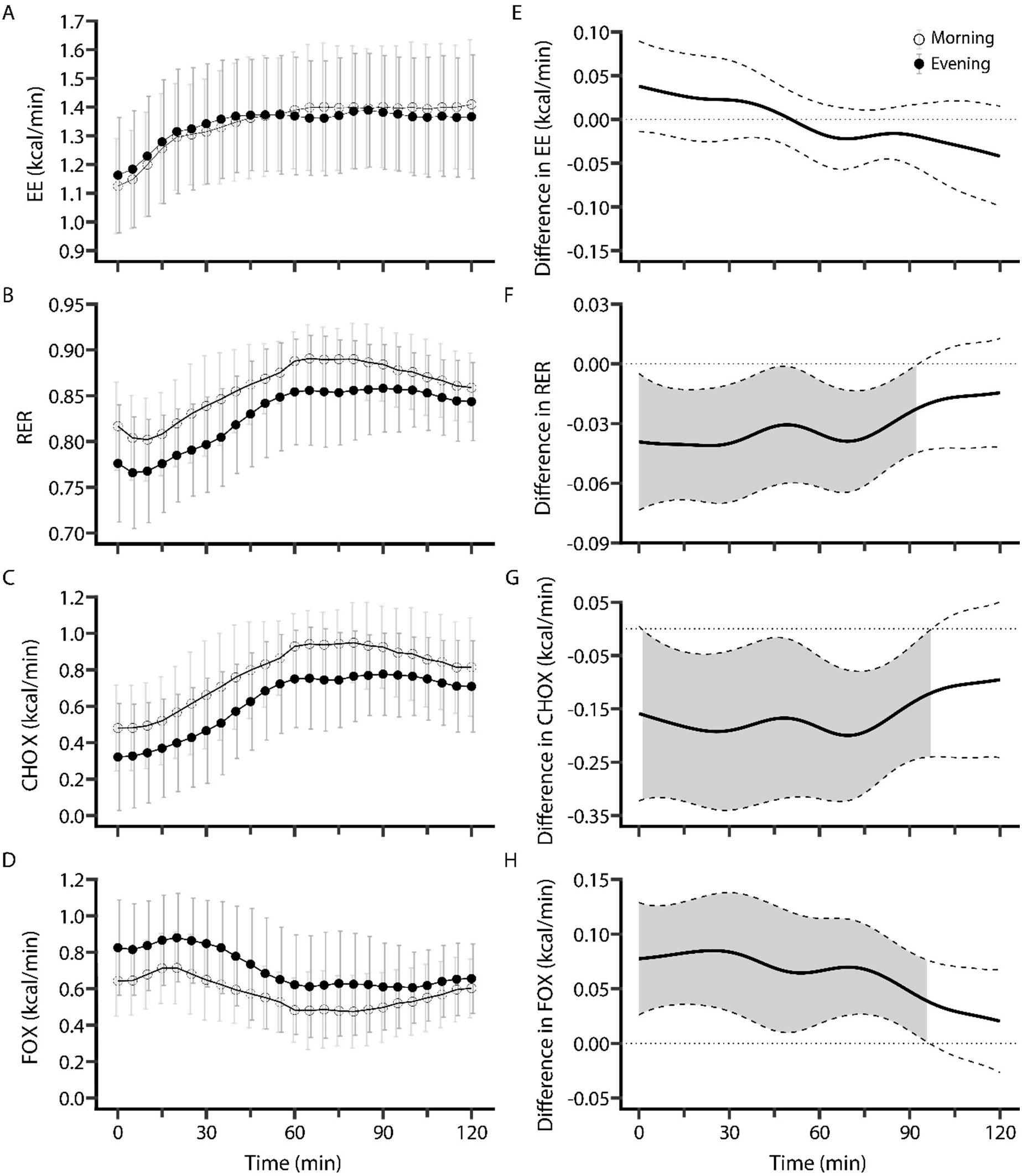
An Evening Meal Increases Fat Oxidation and Decreases Carbohydrate Oxidation. Figure 2 caption: Mean ± SD of EE (A), RER (B), CHOX (C), and FOX (D) during a mixed meal challenge in the morning (open circles) and evening (closed circles) are shown in the left panels. The right panels show the mean difference (evening - morning) as a solid line with the 95% CI as dashed lines between conditions for EE (E), RER (F), CHOX (G), and FOX (H). The dotted lines at zero indicate no difference. Gray shading denotes a 95% CI that does not cross zero, indicating a significant difference between meal conditions at the corresponding timepoint.

### Metabolic Flexibility

To test for differences in metabolic flexibility between morning and evening meals, we repeated functional regression models of each postprandial outcome with an additional covariate for the fasting outcome. In these models, there was no significant effect of time-of-day on postprandial RER (p=0.176) or FOX (p=0.336), indicating no difference in metabolic flexibility between morning and evening meals (**Figure 3**). While there was a significant main effect for EE (p=0.022), none of the 95% CIs excluded the null (**Figure 3A**), indicating insufficient evidence for an effect of time-of-day on changes in postprandial EE by time-of-day. The difference in CHOX from fasting to postprandial was significantly lower in the evening from 70-85 minutes postprandial (p<0.001; −0.12 ± 0.01 kcal/min). To facilitate comparison with prior literature, we also calculated metabolic flexibility as the difference between the peak and nadir of RER divided by the difference in time between them [49]. In these analyses, there was no effect of time-of-day on delta REE (p=0.498), delta RER (p=0.266), delta CHOX (p=0.376), or delta FOX (p=0.935) via paired t-test (**Figure S1**).

**Figure 3.**
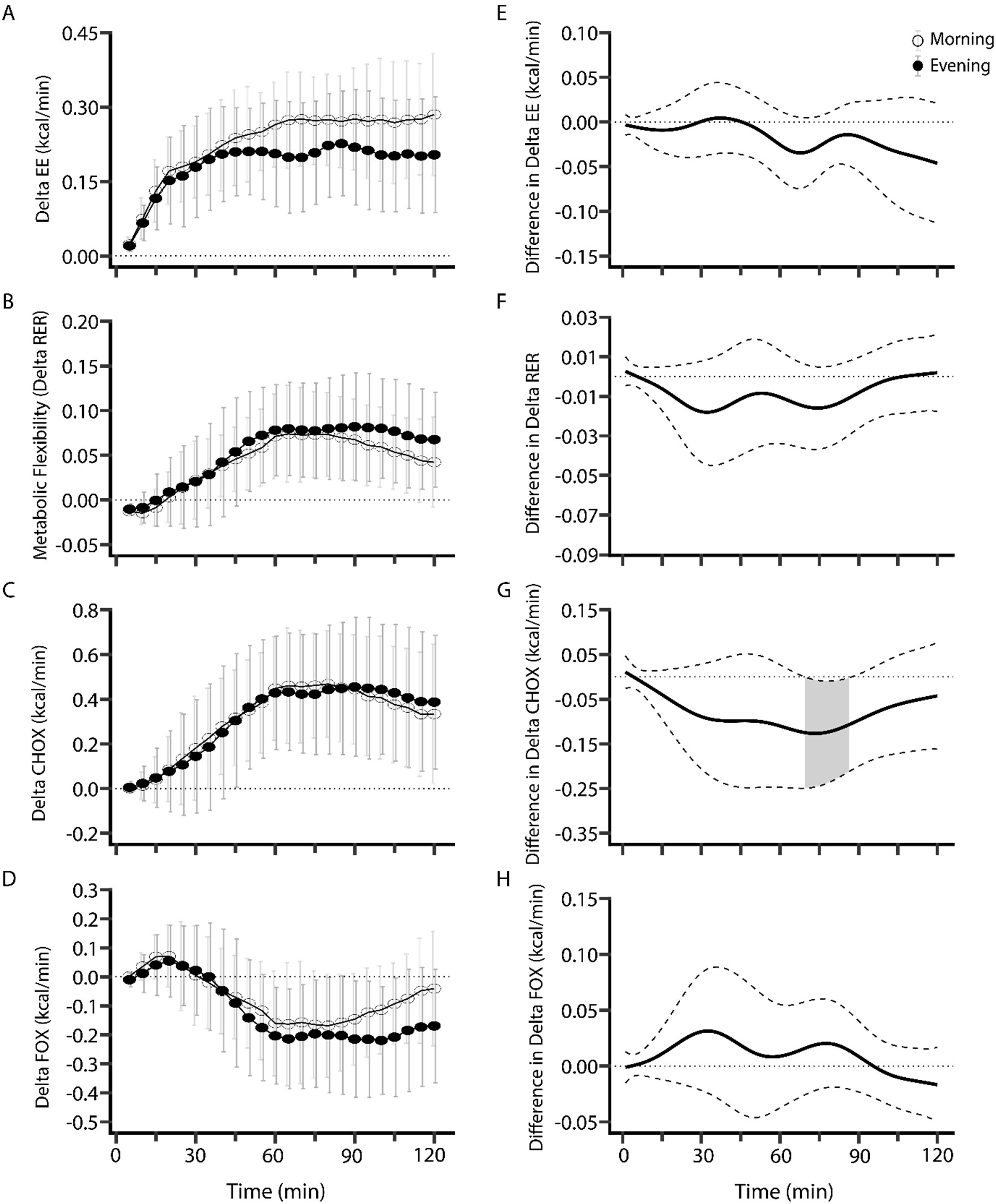
Postprandial Metabolic Flexibility is Not Different Between Morning and Evening Meals. Figure 3 caption: Mean ± SD of the delta from fasting to postprandial for EE (A), RER (B), CHOX (C), and FOX (D) during a mixed meal challenge in the morning (open circles) and evening (closed circles) are shown in the left panels. The right panels show the mean difference between conditions (evening - morning) with a solid line and the 95% CI of the mean difference in dashed lines for EE (E), RER (F), CHOX (G), and FOX (H), each adjusted for the fasting value at the given time of day. The dotted lines at zero indicate no difference. Gray shading denotes a 95% CI that does not cross zero, indicating a significant difference between meal conditions at the corresponding timepoint.

### Effect of Self-Reported Chronotype on Fasting Metabolism

We also conducted an exploratory analysis to understand how chronotype may differentially affect our metabolic outcomes using CMQ score as a self-reported measure of chronotype. In mixed effects models of fasting substrate metabolism, there were no significant chronotype*meal condition interactions for fasting EE (p=0.835), RER (p=0.285), FOX (p=0.396), or CHOX (p=0.490). In subsequent main effects models, there was no effect of chronotype on fasting EE (p=0.524), RER (p=0.562), FOX (p=0.910), or CHOX (p=0.500). After adjustment for chronotype, fasting RER was still lower in the evening (mean difference ± SE (95% CI): −0.04 ± 0.02 (−0.08, −0.001); p=0.043) with higher fasting FOX (0.182 ± 0.064 kcal/min (0.048, 0.032); p=0.011) but without significant changes in CHOX (−0.159 ± 0.087 kcal/min (−0.342, 0.024); p=0.084) or EE (0.04 ± 0.03 kcal/min (−0.02, 0.09); p=0.171).

### Effect of Self-Reported Chronotype on Postprandial Metabolism

We further used function regression models with a chronotype*meal condition interaction to understand how chronotype may impact postprandial substrate metabolism. To simplify this exploratory analysis, we limited these analyses solely to RER as our outcome variable. We compared interaction models with a linear and nonlinear effect of chronotype against their main effects models via F-test. For both linear and nonlinear models, an interaction model was a better fit to the data and improved adjusted R^2^ (both p<0.001; linear model R^2^: 0.690 to 0.706; nonlinear model R^2^: 0.690 to 0.804). Further, the nonlinear interaction model was a better fit to the data than the linear interaction model (p<0.001) and reduced Akaike information criterion (AIC) (7204 to 5550). In the nonlinear interaction model, there was a significant meal condition*chronotype interaction effect (p<0.001) (**Figure 3A**). A more morning chronotype (i.e., a higher CMQ score) is associated with lower fasting RER in the evening and higher postprandial RER in the morning. An intermediate chronotype is associated with little difference between morning and evening meals. A more evening chronotype is associated with a delayed increase in postprandial RER in the evening compared to the evening meal for other chronotypes. To assess differences in metabolic flexibility by chronotype, we adjusted our nonlinear chronotype interaction model for fasting RER (**Figure 3B**). In these models, chronotype impacted evening metabolic flexibility with little difference in morning metabolic flexibility. Morning and intermediate chronotypes displayed appropriate metabolic flexibility in the evening. However, the increase in postprandial RER was delayed in evening chronotypes and did not reach the postprandial levels of morning and intermediate chronotypes until ∼80 minutes postprandial.

### Cardiovascular Outcomes

Brachial DBP (mean difference ± SE: 1.9 ± 0.8 mmHg (0.3, 3.5); p = 0.022), central DBP (1.8 ± 0.9 mmHg (0.1, 3.5); p = 0.035), SEVR (mean difference ± SE: 9.8 ± 3.4; p=0.005), and central pressure AUC during diastole (mean difference ± SE: 140 ± 43; p=0.002) were higher in the evening than the morning (**Table 2**). There was a significant meal condition*time interaction for Pf (p=0.027). Pairwise differences indicate Pf was lower at 60 minutes postprandial in the evening vs. the morning (mean difference ± SE: −3.8 ± 1.2; p = 0.001) without differences at fasting (p = 0.680) or 120-minutes postprandial (p=0.697). No other cardiovascular outcomes differed between the morning and evening meal challenges (**Table 2**). Pairwise contrasts of significant main effects over time during the MMC are presented in Table S1.

**Table 2.** Cardiovascular Outcomes.

| Outcome Measure | Meal Condition | Fasting | 60 min. | 120 min. | P values from Mixed Effects Models |  |  |
|---|---|---|---|---|---|---|---|
|  |  |  |  |  | Time | Meal Condition | Interaction |
| AIx (%) | Morning | 14.4 ± 10.6 | 1.0 ± 9.9 | 7.5 ± 10.9 | **<0.001** | 0.938 | 0.266 |
|  | Night | 12 ± 6.1 | 4.1 ± 11.1 | 6.5 ± 10.5 |  |  |  |
| AIx75 (%) | Morning | 7.9 ± 12.5 | -0.1 ± 10.4 | 5.1 ± 10.8 | **0.005** | 0.604 | 0.398 |
|  | Night | 5.7 ± 8.8 | 1.9 ± 11.4 | 3.1 ± 12.7 |  |  |  |
| Brachial SBP (mmHg) | Morning | 115 ± 8 | 121 ± 11 | 119 ± 13 | **0.023** | 0.995 | 0.313 |
|  | Night | 116 ± 10 | 118 ± 11 | 120 ± 11 |  |  |  |
| Brachial DBP (mmHg) | Morning | 66 ± 8 | 68 ± 7 | 68 ± 9 | 0.147 | **0.022** | 0.847 |
|  | Night | 68 ± 7 | 70 ± 10 | 70 ± 7 |  |  |  |
| Central SBP (mmHg) | Morning | 101 ± 7 | 103 ± 10 | 103 ± 12 | 0.491 | 0.543 | 0.633 |
|  | Night | 103 ± 9 | 102 ± 11 | 105 ± 10 |  |  |  |
| Central DBP (mmHg) | Morning | 67 ± 8 | 70 ± 8 | 70 ± 9 | **0.021** | **0.035** | 0.857 |
|  | Night | 69 ± 8 | 71 ± 10 | 71 ± 7 |  |  |  |
| Pulse Pressure Amplification (%) | Morning | 71 ± 4 | 62 ± 6 | 65 ± 5 | **<0.001** | 0.556 | 0.310 |
|  | Night | 70 ± 3 | 64 ± 6 | 66 ± 4 |  |  |  |
| Central MAP during systole (mmHg) | Morning | 92 ± 7 | 94 ± 9 | 94 ± 11 | 0.304 | 0.273 | 0.761 |
|  | Night | 94 ± 9 | 95 ± 10 | 96 ± 9 |  |  |  |
| Central MAP during diastole (mmHg) | Morning | 74 ± 8 | 76 ± 9 | 77 ± 10 | **0.016** | 0.083 | 0.933 |
|  | Night | 76 ± 8 | 78 ± 10 | 79 ± 9 |  |  |  |
| HR (bpm) | Morning | 61 ± 11 | 73 ± 12 | 70 ± 11 | **<0.001** | 0.222 | 0.633 |
|  | Night | 62 ± 11 | 70 ± 10 | 68 ± 9 |  |  |  |
| PWV (m/s) | Morning | 5.4 ± 0.9 | 6.0 ± 2.2 | 5.5 ± 1.0 | 0.163 | 0.882 | 0.143 |
|  | Night | 5.4 ± 0.7 | 5.4 ± 0.7 | 5.4 ± 0.8 |  |  |  |
| Pf Amplitude (mmHg) | Morning | 28 ± 4 | 30 ± 5 | 28 ± 4 | N/A | N/A | **0.027** |
|  | Night | 27 ± 3 | 27 ± 3 | 28 ± 4 |  |  |  |
| Pb Amplitude (mmHg) | Morning | 13 ± 3 | 12 ± 3 | 13 ± 2 | **0.006** | 0.408 | 0.863 |
|  | Night | 13 ± 2 | 11 ± 2 | 12 ± 2 |  |  |  |
| Ref. Mag. (pb/pf, %) | Morning | 46 ± 7 | 43 ± 18 | 45 ± 6 | 0.351 | 0.817 | 0.728 |
|  | Night | 47 ± 7 | 43 ± 7 | 43 ± 7 |  |  |  |
| Pressure AUC during systole (mmHg*s/min) | Morning | 1994 ± 328 | 2339 ± 344 | 2205 ± 333 | **<0.001** | 0.083 | 0.435 |
|  | Night | 1989 ± 348 | 2210 ± 336 | 2134 ± 315 |  |  |  |
| Pressure AUC during diastole(mmHg*s/min) | Morning | 2824 ± 326 | 2684 ± 426 | 2824 ± 491 | **0.032** | **0.002** | 0.899 |
|  | Night | 2939 ± 323 | 2848 ± 381 | 2963 ± 324 |  |  |  |
| SEVR | Morning | 146 ± 31 | 117 ± 27 | 131 ± 34 | **<0.001** | **0.005** | 0.717 |
|  | Night | 152 ± 31 | 131 ± 21 | 141 ± 20 |  |  |  |

All data are raw values presented as mean ± SD. P-values for time and meal condition are the main effects from a mixed effects model without a time\*meal condition interaction.

## Discussion

In our sample of otherwise healthy young adults, we show substrate utilization in the fasting and postprandial states reflects expected circadian trajectories; FOX was higher and CHOX and RER were lower in the evening compared to the morning without differences in metabolic flexibility to a mixed meal. We further accounted for inter-individual differences in preference for morning activity as a proxy for chronotype, showing greater morningness is associated with greater fasting and postprandial FOX. Diastolic blood pressure was elevated in the evening compared to the morning without differences in the postprandial blood pressure response. Contrary to our hypotheses, arterial stiffness via cf-PWV was not different between morning and evening. In our sample of young adults free of overt pathology, time-of-day differences in cardiometabolic responses to feeding attenuate and are no longer meaningful when accounting for circadian variation in fasting physiology, though the magnitude of this difference may be influenced by chronotype.

We observed higher evening FOX, lower RER, and lower CHOX in the fasting and postprandial states, which align with other reported time of day changes in postprandial substrate oxidation [9, 50]. These differences persisted through ∼90 minutes postprandial, likely reflecting slower glucose uptake in the evening [51]. The postprandial discrepancy between morning and evening CHOX was larger than the postprandial increase in FOX (−0.15 kcal/min vs. +0.07 kcal/min; **Figure 2G-H**), suggesting the observed changes in whole-body metabolism may be due to lower insulin sensitivity [52, 53] rather than increased endogenous fatty acid oxidation [54]. Because skeletal muscle mitochondrial oxidative capacity *in vivo* peaks in the late evening (∼23:00) [55], the observed morning-evening differences may be even greater with a later meal.

By having participants fast for ten hours prior to each meal, we were able to isolate the underlying time-of-day effects from the confounding metabolic effects of a previous meals [56]. Further, our use of functional regression and adjustment for fasting RER do not limit us to a single postprandial timepoint, which overlooks the dynamic nature of postprandial metabolism. Contrary to our hypothesis, the ability to appropriately increase RER in response to a MMC was not affected by time of day, indicating similarly appropriate metabolic flexibility in the morning and evening (**Figure 3B**). This is contrasted by a recent study by Dörner et al. [9], who showed a decrease in the change in RER in response to successive mixed meals in eight young adults. Participants in Dörner et al. fasted for six hours between meals, which is shorter than the ten hours between meals for our participants. Our use of morning vs. evening meals with an equal pre-meal fasting period differs from their approach of three successive meals, making it difficult to directly compare our findings.

By adjusting our models for the fasting outcome measured prior to each meal (i.e., in the morning and evening), we also show a small effect of time-of-day on postprandial CHOX with no effect on postprandial FOX, RER, or EE. Because the effect of time-of-day on the postprandial rise in CHOX is small (0.12 kcal/min) and only significant from 75-90 minutes, this difference has questionable physiological relevance. Combined with our null findings for RER, EE, and FOX, the observed postprandial effects are likely due to the metabolic effects of the underlying circadian rhythm and not a unique time-of-day effect on postprandial substrate oxidation. Bo et al. also compared morning and evening feeding of a single, solid meal in 20 young adults with a similar pre-meal fasting duration [50]. When studying substrate utilization in the later postprandial period (i.e., 120-180 minutes post-meal), they observed greater fasting to feeding changes in RER and an increase in FOX following an evening meal. Importantly, the fasting-feeding difference in FOX and CHOX was not different by time of day, suggesting differences in fasting substrate utilization by time of day drove observed postprandial differences, in agreement with our findings. While Bo et al. observed greater postprandial EE in the morning than in the evening, this is likely due to their larger meal (1170 kcal vs. our 635 ± 118 kcal) and greater energy from protein (30% vs. our 18%), leading to a greater thermic effect of feeding [57]. When a meal of a similar size was used, there were no differences in thermic effect of feeding between morning and evening meals [9, 50, 58]. The use of stable isotope tracers would allow us to better characterize ingested nutrient fates [59, 60], which can remain in circulation or be taken up by peripheral tissues for oxidation or storage [61]. Likewise, given the relevance of insulin and peripheral insulin sensitivity to postprandial substrate utilization [62], future work should investigate time-of-day differences in postprandial metabolism in populations with worse overall insulin resistance to explore differences in time-of-day feeding responses by disease status. Indeed, the diurnal rhythmicity of the serum and adipose tissue metabolomes differ between adults with type 2 diabetes compared to healthy controls [63].

We further explored the potential for self-reported chronotype to modulate postprandial metabolism. Interestingly, time-of-day differences in modeled RER were largely non-existent with intermediate chronotypes. In earlier chronotypes, fasting RER was unexpectedly higher in the morning and lower in the evening with similar postprandial RER in the later portion of the MMC. In later chronotypes, fasting RER was similar in the morning and evening, but evening increases in postprandial RER were delayed compared to the morning. In analyses adjusted for fasting RER, chronotype does not impact metabolic flexibility to a morning meal (**Figure 4B**). However, an evening chronotype is associated with delayed metabolic flexibility in the evening, where modeled RER adjusted for fasting RER (i.e., the change in RER from fasting to feeding) does not rise above 0.90 until 100 minutes postprandial. This is contrasted by morning and intermediate chronotypes, whose modeled RER rises quickly and remains elevated through 120 minutes. Collectively, these data indicate greater variation in fasting and postprandial substrate metabolism by time of day in more extreme chronotypes, with the greatest differences by chronotype in the early evening.

**Figure 4.**
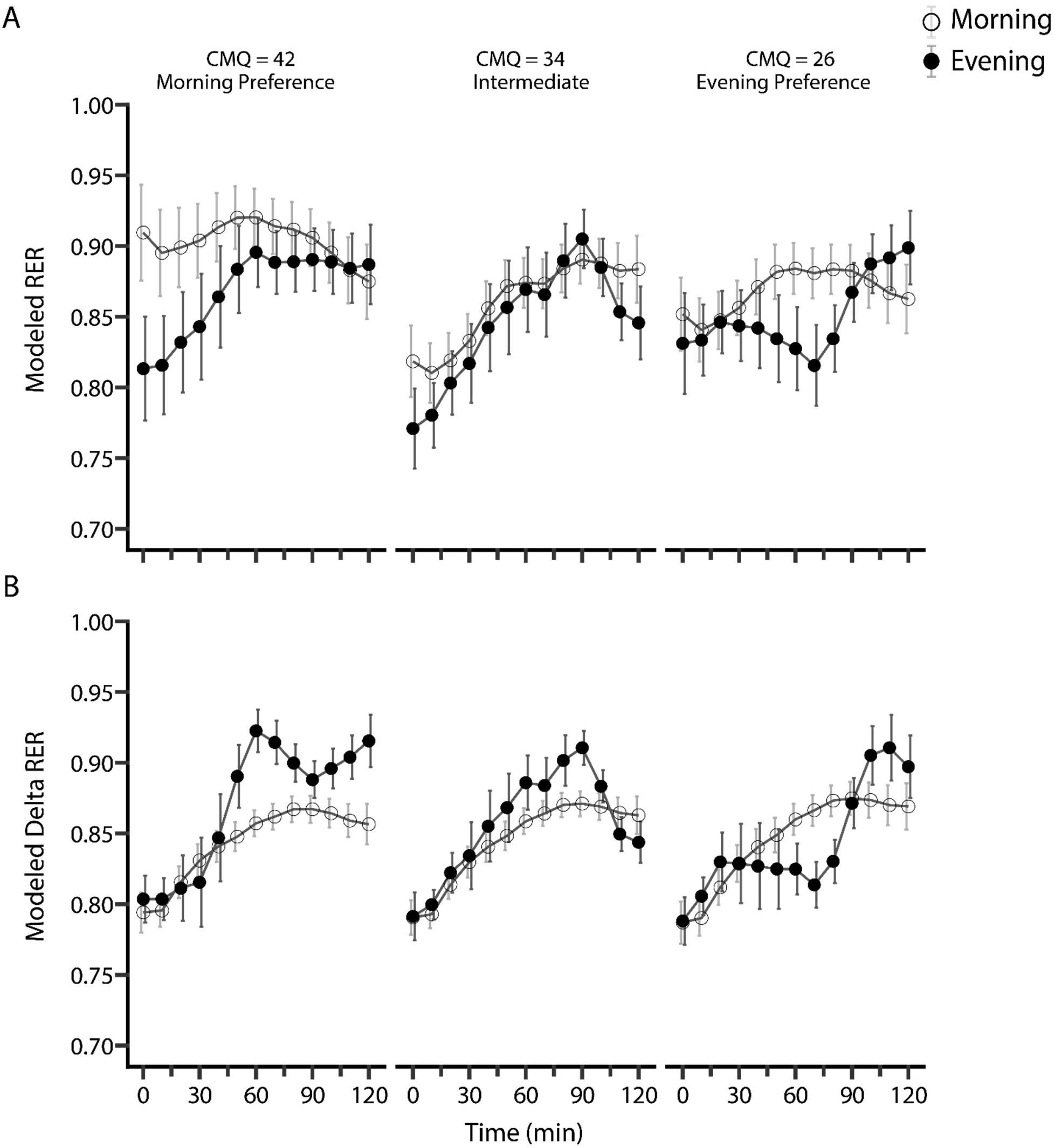
More Extreme Chronotypes Have Greater Differences Between Morning and Evening Substrate Metabolism. Figure 4 caption: Data are presented as modeled mean ± SE for morning (open circles) and evening (closed circles) RER at representative values of CMQ in our sample (mean: 34; SD: 8), showing an association between more extreme chronotypes and larger morning-to-evening differences in RER (A). Metabolic flexibility is delayed in more evening chronotypes. Respiratory exchange ratio (RER).

Few experimental studies have examined chronotype as a source of inter-individual variability. In a crossover trial of routine dinner (18:00) vs. late dinner (22:00) with H_2_^18^O and [^2^H_31_]-palmitate, early habitual bedtime was associated with decreased dietary FOX at late dinner in healthy young adults [64]. In a crossover trial of high glycemic index meals in the morning (07:00) vs. evening (20:00), morning chronotypes had greater 2-hour postprandial blood glucose via continuous glucose monitor in the evening while evening chronotypes showed no difference in glycemic response by time of day [23]. Of note, the 40 participants were selected as those with the most extreme chronotypes (i.e., the highest and lowest questionnaire scores) among an eligible sample of 231 participants, which limits comparison with our study of a narrower range of chronotype scores. Future studies, especially those conducting trials of different nutritional interventions across the day, should consider exploring outcomes by chronotype to better clarify its contribution to inter-individual variability in substrate metabolism.

Our observed differences in substrate oxidation by chronotype occurred despite the modeled values of CMQ (e.g., 26, 34, 42) all falling within the intermediate chronotype range (i.e., 23-43) [25], which is the most common chronotype in the population [19]. We expect these differences would be even larger in true morning or evening chronotypes. While we did not recruit participants from the full range of chronotype in the population, we believe these findings highlight the importance of considering chronotype when examining the circadian effects of metabolism and may underlie the metabolic deficiencies often observed in evening chronotypes [15]. We did not power our study for interactions by chronotype and chose to model chronotype nonlinearly based on improved model fit rather than an *a priori* justification; further studies will be needed to validate our observed nonlinear effect of chronotype.

The meal times employed in this study reflect mealtimes characteristic of earlier chronotypes [65], which may have affected postprandial substrate utilization when considered in relation to habitual intake timing [66]. Because we did not standardize mealtimes to a circadian rhythm, the breakfast consumed prior to the evening MMC may be disproportionately early for evening chronotypes. We were also unable to standardize energy balance in the 24 hours prior to each MMC, such that participants fasted overnight before eating breakfast and fasting during the day prior to the evening MMC. Nevertheless, we show an association between self-reported chronotype and postprandial substrate oxidation which may underlie the metabolic differences by chronotype.

In concert with these metabolic responses, vascular responses to feeding were largely appropriate but not different by time of day. AIx and AIx75 were reduced at one-hour postprandial, which was also observed in a previous study in young adults and is likely the result of insulin-mediated vasodilation [67]. Likewise, heart rate increased, likely to facilitate digestion and delivery of macronutrients to peripheral tissues. Time-of-day differences in fasting hemodynamics were modest, though we did observe an increase in central and brachial diastolic blood pressure from morning to evening. Our finding of elevated blood pressure in the evening is consistent with several studies [6, 7, 68, 69], including one which also employed daytime fasting after a morning meal [70].

Taken together, metabolic flexibility to morning and evening MMCs was similar, and differences in postprandial substrate oxidation between morning and evening were largely explained by greater fasting FOX and reduced fasting CHOX (resulting in lower RER), regardless of morningness and without significant influence on hemodynamic responses to feeding. Although previous studies have demonstrated time-of-day dependent changes in postprandial substrate oxidation [9, 50, 58], our use of a matched pre-meal fasting duration allows us to isolate the contributions of a single meal to early postprandial responses to feeding. Further, the caloric content of our mixed meal is largely representative of lunch energy content for the average U.S. adult [71], enhancing the practical relevance of our study design compared to studies using much larger meals [50]. Our statistical approach protects against inflated type I errors over the frequently sampled postprandial period while still offering greater time-course resolution needed to understand the dynamic postprandial state. Furthermore, we investigated the potential of chronotype to modulate postprandial substrate dynamics. Our approach of expressing chronotype as a continuous rather than categorical variable better aligns with the true physiological spread of behavior throughout the day [19]. Collectively, our approach allowed us to uncover heterogeneity in differences between morning and evening fasting and postprandial substrate oxidation, highlighting the importance of maintaining the continuous nature of this variable.

Understanding evening feeding is necessary to properly characterize metabolic nutrient handling by time-of-day and its purported role in metabolic dysfunction. Since dinner compromised 35% of daily caloric intake and over 70% of U.S. young adults report late-night eating [71], it is imperative to conduct further studies in the evening. Likewise, since later chronotypes are associated with worse cardiometabolic outcomes, studying metabolic responses in this population during their habitual feeding window is most ecologically relevant. Here we have shown that while the metabolic and vascular responses to feeding are similar overall between morning and evening when accounting for observed differences in fasting metabolism, they vary by self-reported chronotype.

## Data Availability

All data produced in the present study are available upon reasonable request to the authors

Upon publication in a peer-reviewed journal, all analysis files and code will be deposited into a publicly available repository. Until then, data are available via the corresponding author upon reasonable request.

## Acknowledgements

We thank the participants for volunteering their time and efforts for this study. We also acknowledge the use of BioRender to create Figure 1 (Foreman, N. (2026); https://BioRender.com/aworf3q). The current affiliation for EUF and NAF is National Institute of Diabetes Digestive and Kidney Diseases, National Institutes of Health, Bethesda, MD.

## Author Contributions

EUF, NAF, and MDB conceived and designed the experiments; EUF, NAF, SS, EG, and AN performed experiments; NAF and AC analyzed data; EUF, NAF, LNW, and MDB interpreted results; NAF prepared figures; EUF and NAF drafted the manuscript; EUF, NAF, AC, LNW, and MDB edited and revised the manuscript; EUF, NAF, SS, EG, AN, AC, LNW, and MDB approved the final version of the manuscript.

## Supplemental Materials

### Graded Exercise Testing

A graded exercise test on a cycle ergometer (Monark LC6 novo; Varberg, Sweden) with breath-by-breath indirect calorimetry (Cosmed Quark CPET; Rome, Italy) was used to measure VO_2MAX_. Heart rate was monitored via a chest-strap heart rate monitor (Polar H10; Kempele, Finland). After five minutes of seated rest, participants cycled for three minutes each at 25, 50, and 75W before increasing at 25W/min to volitional exhaustion or inability to maintain ≥ 50 revolutions per minute. The highest 30-second value was taken as VO_2MAX_.

### Indirect Calorimetry Data Processing

For fasting and postprandial indirect calorimetry (IC), we removed all data for 30 seconds following a calibration event, which occurred every five minutes, and from the first three minutes of each measurement window. We removed data surrounding data calibration events because they were associated with large, supraphysiologic spikes in RER and created artificial noise in the data. Fasting indirect calorimetry (IC) data were analyzed using a six-minute rolling average (i.e., n=72) via zoo::rollmeanr. We chose to use a six-minute window as this minimized the intra-individual day-to-day variability in fasting RER in a similar population from our lab (n=18, data not yet published) compared to other rolling average options ranging from 0.25 to 12 minutes. We selected the minimum RER and EE values as fasting RER and EE, respectively. We selected CHOX and FAO at the time of minimum RER as fasting CHOX and FAO.

For postprandial IC, we used the ventilatory_outliers function within the gasExchangeR package, a publicly available R package, to remove outliers in VO_2_ and VCO_2_ at ± 2.5 SD and for each IC outcome variable at ± 3 SD. To smooth data, we calculated a 6-minute rolling average using zoo::rollmeanr for each measurement and summarized data into 60-second bins (i.e., 0-1 min. becomes 1, 1-2 min. becomes 2, etc.). Because functional regression requires complete data, we imputed each outcome variable from fasting until 120 minutes. Prior to imputation, we extended the fasting value for each outcome variable by 15 minutes and added random noise of mean zero and SD equal to 0.3% of the fasting value for each outcome variable to better account for the steady-state nature of fasting metabolism; this resulted in IC data from −15 to 120 minutes. We used stats::splinefun with method=“natural” to impute IC data and visually compared imputed and measured data to verify accuracy before removing the fasting data with random noise. All IC data were centered and scaled prior to analysis via functional regression.

### Functional Regression

We use function-on-scalar regression via the pffr function in the refund package (version 0.1-40) [46]. Here, we fit a scalar-on-function model with outcome Y_i_(*t*) representing the outcome of interest as a function for the i^th^ participant at timepoint t. Predictors take the form of *β_0_(t) + Σ^p^_j=1_ z_i,j_β_j_(t)* for scalar predictors *j* = 1, …, *p*. Each predictor term is defined as follows: *β*_0_(*t*) represents the functional intercept over all *t*; *z*_i,j_ is the measured value for participant *i* and the scalar predictor *j*; and *β*_j_(*t*) is the coefficient function which describes how scalar *j* affects the outcome Y_i_(*t*) across all values of *t*. Here, we fit four functional regression models, one for each whole-body metabolic outcome. In each model, Y_i_(*t*) represents the given outcome measure (i.e., RER, FOX, CHOX, and EE) under both meal conditions with a binary predictor for male sex. We also include a subject-specific random intercept function to account for repeated functional outcome measures. We estimate the coefficient functions using penalized B-splines with k=30, which is larger than necessary to allow for flexibility in estimating the coefficient function. We also include a penalty on the second derivative to enforce smoothness of the estimated coefficient function, where k is the number of basis functions. We calculate simultaneous 95% confidence intervals (CI) to determine the effect of diet on each metabolic outcome. Importantly, this does not increase Type I error rate by testing for differences by diet individually across all 120 observed timepoints, and smaller differences can be detected compared to a mixed effects model with a Bonferroni-corrected alpha level of 0.000416 (i.e., 0.05/120). Each point where the 95% CI does not cross zero indicates the variable of interest has a significant effect on the outcome at that point in time. To visualize our results, we present raw data for each outcome as mean ± SD for each condition along with panels showing each pairwise difference across conditions. Colored shading shows where the 95% CI does not cross zero. In interaction models, we present data as mean ± SE.

---

**Figure S1:**
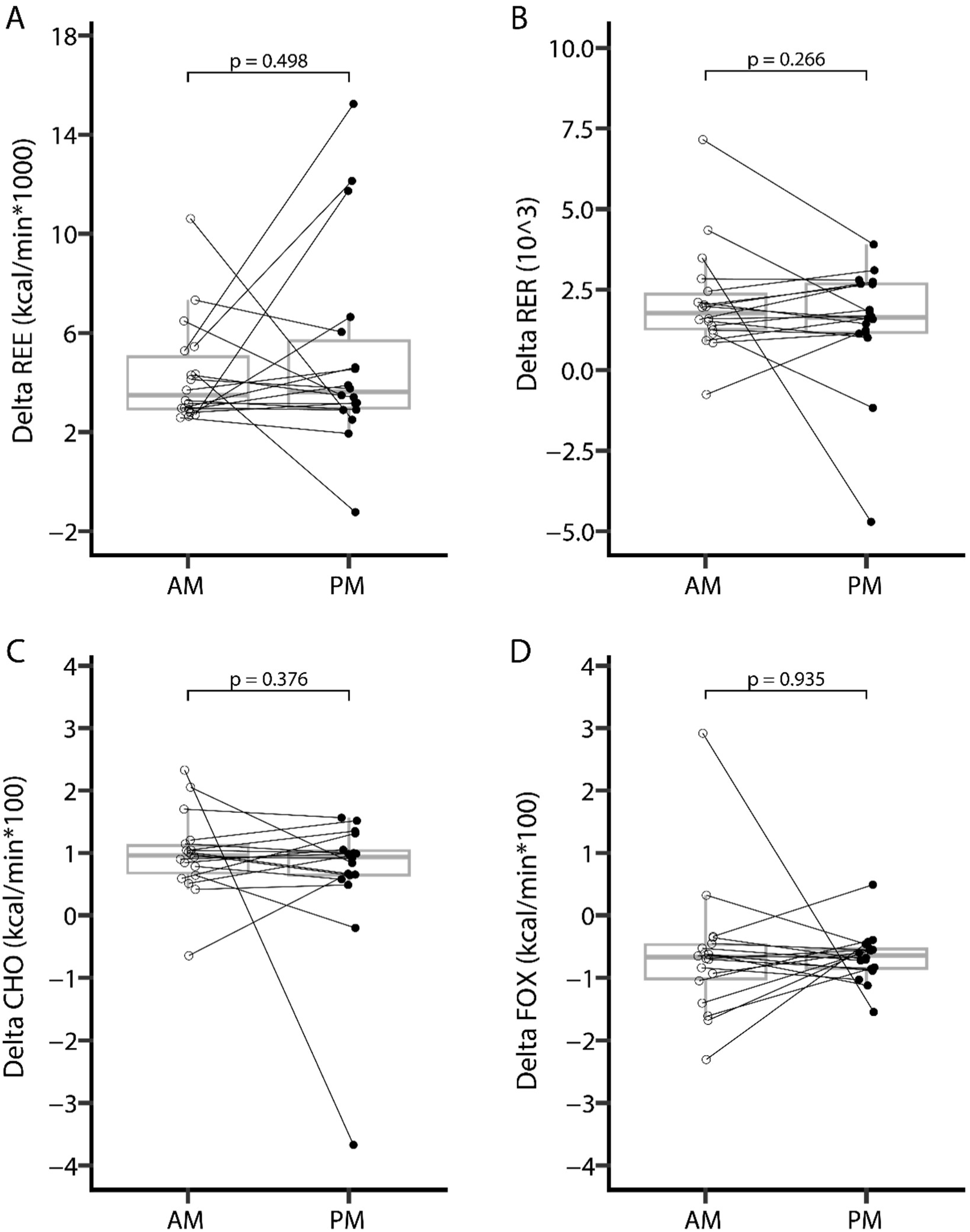
Metabolic Flexibility Does Not Differ Between Morning and Evening Meals When Calculated Using Conventional Methods. Metabolic flexibility calculated as the difference between the peak and nadir of each outcome divided by the difference in time between them [49] showed no effect of time-of-day on delta REE (A), delta RER (B), delta CHO (C), or delta FOX (D) via paired t-test. Resting energy expenditure (REE); Respiratory exchange ratio (RER); Carbohydrate oxidation (CHO); Fat oxidation (FOX).

**Table S1.** Pairwise Comparisons of Significant Main Effects of Time During the Mixed Meal Challenge.

| Outcome Variable | Pairwise Comparison |  | Adjusted P-value | Estimate | SE | Lower CI | Upper CI |
|---|---|---|---|---|---|---|---|
| AIx (%) | 0 | 60 | **<0.001** | 10.6667 | 1.7404 | 6.4181 | 14.9152 |
|  | 0 | 120 | **<0.001** | 6.2222 | 1.7404 | 1.9737 | 10.4708 |
|  | 60 | 120 | **0.012** | -4.4444 | 1.7404 | -8.693 | -0.1959 |
| AIx75 (%) | 0 | 60 | **0.004** | 5.8611 | 1.744 | 1.6038 | 10.1184 |
|  | 0 | 120 | 0.126 | 2.6944 | 1.744 | -1.5628 | 6.9517 |
|  | 60 | 120 | 0.109 | -3.1667 | 1.744 | -7.4239 | 1.0906 |
| Brachial SBP | 0 | 60 | **0.027** | -0.0333 | 0.0138 | -0.067 | -0.00005 |
|  | 0 | 120 | **0.027** | -0.0338 | 0.0138 | -0.0675 | -0.00001 |
|  | 60 | 120 | 0.970 | -0.0005 | 0.0138 | -0.0342 | 0.0332 |
| Brachial MAP | 0 | 60 | **0.023** | -2.8333 | 1.1456 | -5.63 | -0.0367 |
|  | 0 | 120 | **0.022** | -3.1389 | 1.1456 | -5.9356 | -0.3422 |
|  | 60 | 120 | 0.790 | -0.3056 | 1.1456 | -3.1022 | 2.4911 |
| Central DBP | 0 | 60 | **0.025** | -2.6111 | 1.0472 | -5.1674 | -0.0548 |
|  | 0 | 120 | **0.025** | -2.5556 | 1.0472 | -5.1118 | 7.00E-04 |
|  | 60 | 120 | 0.9578 | 0.0556 | 1.0472 | -2.5007 | 2.6118 |
| Heart Rate | 0 | 60 | **<0.001** | -10.1111 | 1.4213 | -13.5807 | -6.6416 |
|  | 0 | 120 | **<0.001** | -7.0556 | 1.4213 | -10.5251 | -3.586 |
|  | 60 | 120 | 0.034 | 3.0556 | 1.4213 | -0.414 | 6.5251 |
| Diastolic MAP | 0 | 60 | **0.0496** | -2.4444 | 1.1288 | -5.2001 | 0.3112 |
|  | 0 | 120 | **0.019** | -3.1667 | 1.1288 | -5.9223 | -0.411 |
|  | 60 | 120 | 0.524 | -0.7222 | 1.1288 | -3.4779 | 2.0334 |
| Pb | 0 | 60 | **0.006** | 1.5612 | 0.4899 | 0.36 | 2.7625 |
|  | 0 | 120 | 0.3916 | 0.418 | 0.485 | -0.7712 | 1.6072 |
|  | 60 | 120 | **0.034** | -1.1432 | 0.4899 | -2.3444 | 0.0581 |
| Pulse Pressure Amplification (%) | 0 | 60 | **<0.001** | 6.7948 | 0.8949 | 4.6103 | 8.9793 |
|  | 0 | 120 | **<0.001** | 4.6216 | 0.8949 | 2.4371 | 6.8061 |
|  | 60 | 120 | **0.017** | -2.1732 | 0.8949 | -4.3577 | 0.0113 |
| Diastolic AUC | 0 | 60 | **0.046** | 115.8056 | 52.6332 | -12.68 | 244.2912 |
|  | 0 | 120 | 0.823 | -11.8056 | 52.6332 | -140.2912 | 116.68 |
|  | 60 | 120 | **0.046** | -127.6111 | 52.6332 | -256.0967 | 0.8745 |
| Systolic AUC | 0 | 60 | **<0.001** | -282.5278 | 47.8135 | -399.2476 | -165.8079 |
|  | 0 | 120 | **<0.001** | -177.6667 | 47.8135 | -294.3865 | -60.9468 |
|  | 60 | 120 | **0.031** | 104.8611 | 47.8135 | -11.8587 | 221.581 |
| SEVR | 0 | 60 | **<0.001** | 24.6667 | 4.193 | 14.431 | 34.9023 |
|  | 0 | 120 | **0.005** | 12.7778 | 4.193 | 2.5421 | 23.0135 |
|  | 60 | 120 | **0.006** | -11.8889 | 4.193 | -22.1246 | -1.6532 |

See Table 2 and main text for abbreviations

